# A framework for network-based epidemiological modeling of tuberculosis dynamics using synthetic datasets

**DOI:** 10.1101/2020.03.30.20047795

**Authors:** Marissa Renardy, Denise E. Kirschner

**Affiliations:** University of Michigan, Department of Microbiology and Immunology, Ann Arbor, MI, USA

**Keywords:** tuberculosis, epidemiology, network-based model, synthetic population

## Abstract

We present a framework for discrete network-based modeling of TB epidemiology in US counties using publicly available synthetic datasets. We explore the dynamics of this modeling framework by simulating the hypothetical spread of disease over two years resulting from a single active infection in Washtenaw County, MI. We find that for sufficiently large transmission rates that active transmission outweighs reactivation, disease prevalence is sensitive to the contact weight assigned to transmissions between casual contacts (that is, contacts that do not share a household, workplace, school, or group quarter). Workplace and casual contacts contribute most to active disease transmission, while household, school, and group quarter contacts contribute relatively little. Stochastic features of the model result in significant uncertainty in the predicted number of infections over time, leading to challenges in model calibration and interpretation of model-based predictions. Finally, predicted infections were more localized by household location than would be expected if they were randomly distributed. This modeling framework can be refined in later work to study specific county and multi-county TB epidemics in the US.

## 1 Introduction

Mathematical and computational epidemiological modeling has been widely applied to a variety of diseases. In particular, tuberculosis (TB) has been the focus of much work; see, for example, Blower et al. (1995); Castillo-Chavez and Feng (1998); Lietman and Blower (2000); Ziv et al. (2004); Abu-Raddad et al. (2009); Guzzetta et al. (2011); Tian et al. (2013); Kasaie et al. (2014); Knight et al. (2014); Prats et al. (2016); Renardy and Kirschner (2019). There are many approaches to modeling at an epidemiological scale, including use of ordinary and partial differential equations, stochastic processes, agent-based models, and network structure. In heterogeneous populations, contact networks have been shown to have significant impacts on the spread of infectious diseases (Bansal et al., 2007), and there are many ways to model such networks (Keeling and Eames, 2005). Here, we investigate a novel approach to modeling TB epidemics by creating a framework combining publicly available synthetic data, individual-based network modeling, and the natural history of pulmonary TB.

In this work, we develop a new framework for studying TB epidemics. We create an individual-based network model for TB and explore its capabilities using publicly available synthetic datasets based on census data. In this study, we use a synthetic dataset for Washtenaw County, MI, which was chosen solely because it is where University of Michigan and the authors are located, but this framework could be applied to any US county. A major benefit of this framework is the utilization of publicly available synthetic population datasets described by Wheaton et al. (2009) to create a realistic contact network within a population, as well as socio-demographic attributes for all individuals, based on census data. These synthetic population data have been used in several epidemiological modeling studies for influenza and MRSA within US counties and cities (Lee et al., 2010a,b, 2011; Cooley et al., 2010; Macal et al., 2012; Macal et al., 2014), but have not yet been utilized for modeling TB. A similar type of synthetic population was constructed based on a European study by Merler and Ajelli (2010), and was used to model TB dynamics in the state of Arkansas (Guzzetta et al., 2011). That study showed that a model including socio-demographic features such as households, workplaces, and schools was better able to reproduce epidemiological data than simpler model representations, and therefore better able to make more accurate intervention predictions. However, this dataset is not publicly available, limiting reproducibility. Our model differs from this previous work in that we are presenting a framework that links a network-based model together with publicly available synthetic datasets and can be easily adapted to model any US county, and thus is more reproducible and extendable within the US.

As this paper is focused on building and understanding synthetic datasets in network models, rather than specifically predicting TB outcomes, we do not calibrate the model to epidemiological data within Washtenaw County and thus we make no specific predictions about the spread of TB. Rather, we use randomization and parameter exploration to study general model behavior.

## 2 Biological background

TB is an infectious disease cause by the bacterium <u>Mycobacterium tuberculosis</u> (Mtb). It typically is inhaled and infects the lungs, spreading from person to person through the air, though Mtb can also infect other parts of the body. Most individuals exposed to TB develop a <u>latent</u> infection, meaning that they experience no clinical symptoms and cannot transmit Mtb. A small proportion (roughly 5-10%), on the other hand, develop an <u>active</u> infection soon after exposure (within 2 years) (CDC, Division of Tuberculosis Elimination, 2011). Individuals with a latent infection may remain latent for many years, but may develop an active infection later in life either through reactivation of the original infection or reinfection due to a second exposure. Overall, people infected with Mtb have a 5–10% lifetime risk of developing an active infection (WHO, 2019). This probability is significantly increased for individuals who have additional risk factors such as HIV-1, diabetes, smoking, and alcohol abuse.

TB is the leading cause of death worldwide from a single infectious agent, and roughly a quarter of the world’s population carries a latent TB infection (WHO, 2019). In the US, the incidence of active TB was 2.8 per 100,000 in 2018 (CDC, 2019). While TB incidence has steadily declined in the United States (US) since the 1990s (CDC, 2019), effective strategies for TB elimination continue to be elusive. The CDC has indicated that the current rate of decline in TB incidence in the US is insufficient for reaching elimination targets by the year 2100 (Stewart et al., 2018). Novel intervention strategies are likely needed to meet these goals. While the goal of this paper is not to make specific predictions for TB in Washtenaw County, we are creating a framework that going forward could be applied to specific populations and their corresponding datasets to make more accurate predictions than are currently possible.

## 3 Methods

### 3.1 Network model for TB epidemiology in a synthetic population

Since TB is a low-incidence disease within the US and there are many factors that affect susceptibility, stochastic effects and population heterogeneity are of great importance to understanding disease dynamics. These factors should be explicitly considered in building a mathematical or computational model. Thus, discrete model frameworks such as network-based (NBM) and agent-based models (ABMs) are an ideal choice when sufficient information is available describing demographic features and contact patterns of a population of interest. Socio-demographic data are available for US states and counties via the US Census Bureau. In addition, <u>synthetic population datasets</u> have been created from these data for use in agent-based models (Wheaton, 2014; Wheaton et al., 2009). These synthetic population datasets consist of information on individuals together with demographic, geographical, and socioeconomic features that are assigned to households, workplaces, schools, and group quarters in a way that is consistent US Census data. Synthetic datasets allow establishment of a realistic contact network in which diseases can spread. These contact networks may be paired with an epidemiological model to allow for tracking of key features such as location of transmission events and demographic information of the infected population.

Using synthetic populations, models of TB epidemiology can be studied at a variety of spatial scales ranging from groups of states to single counties depending on a population of interest and computational resources available. We focus here on a single county level, which allows for intervention strategies to be designed and optimized for specific local populations. This can reveal differences between intervention efficacies in different locales and subpopulations, and identify location-specific high-risk groups. This approach will be translatable to future models that are specifically targeting a population.

Of the existing publicly available epidemic modeling frameworks, this model is similar in concept to FRED (Grefenstette et al., 2013) and EpiSimS (Mniszewski et al., 2008), which use synthetic population datasets to model flu-like epidemics. The novelty of the model presented here is the application of synthetic population datasets to model TB epidemiology, which has multiple paths to active infection and does not necessarily follow the traditional SEIR framework (see Section 3.1.2 for a description of disease progression).

#### 3.1.1 Population and contact networks

To explore the use of synthetic population data in studying TB, we chose to develop a network-based model (NBM) capturing epidemiological dynamics of TB within a human population. We base the model after a set of models that we have developed previously using systems of ordinary differential equations (ODE), an ABM formulation, and an age-structured partial differential equation (PDE) formulation (Guzzetta et al., 2011; Renardy and Kirschner, 2019). Our network model population is based on synthetic population data for Washtenaw County, MI taken from Wheaton (2014). Synthetic datasets are available through RTI International at both the state and county levels for all US states and counties. These datasets can be accessed at https://fred.publichealth.pitt.edu/syn_pops. The population is comprised of individuals with socio-demographic attributes such as age, sex, race, and household income. Individuals are assigned to geospatially explicit households, workplaces, schools, and/or group quarters, such as college dorms, prisons, and nursing homes, in a way that is consistent with available census data. The methodology for creating these synthetic datasets is described in Wheaton et al. (2009).

The synthetic population for Washtenaw County, MI includes 343,322 individuals, 137,181 households, 29,291 workplaces, 109 schools, and 47 group quarters. 16,502 of the individuals in Washtenaw County reside in group quarters. Of the 47 group quarters, 20 are college dorms (containing a total of 13,873 individuals), 14 are prisons (containing a total of 1,944 individuals), and 13 are nursing homes (containing a total of 685 individuals). The median school size is 571 students, and the median workplace size is 5 workers. The largest workplace employs 25,358 individuals. The age distribution of the population is: 12% children under 10, 13% 10–20 years old, 19% 20–30 years old, 13% 30–40 years old, 14% 40–50 years old, 14% 50–60 years old, and 15% over age 60. Since other socio-demographic factors such as sex, race, and income do not affect disease transmission in our current model, we will not include that information here.

The synthetic population datasets contain, for each individual, their socio-demographic features as well as unique identifiers for their household, workplace, school, and/or group quarter. In addition, the datasets contain locations and descriptive attributes of each household, workplace, school, and group quarter, which can be linked with the people data via the unique identifiers. Thus, it can be easily identified which individuals share a household, workplace, school, or group quarter.

While the synthetic population datasets do contain geospatial coordinates for each of the locations, in our NBM agent interactions are simulated based on contact networks and not physical locations. Agents interact with one another via households, workplaces, school, group quarters, and casual/random contacts. Different types of contacts are assigned different contact weights, with larger contact weights representing more frequent and/or more prolonged contact; e.g., a household contact may be weighted heavier than a school contact, which may be weighted heavier than a casual contact. Agents in large schools/workplaces (more than 50 people) are assumed to have regular contact with at least 10 and no more than 50 other members of their school/workplace. The upper bound of 50 contacts was chosen arbitrarily to avoid excessive and unrealistic transmission within workplaces. If data on the appropriate number of contacts were available, this upper bound could be modified. The number of school/workplace contacts for an individual is chosen randomly, and the contacts themselves are also chosen randomly among all other individuals in that school/workplace.

To simulate casual contacts, all agents not in prisons are randomly assigned to have contact with 10–50 other agents in the population. This represents all contacts that do not share a household, workplace, school, or group quarter. Since we are modeling a population within a single county, we assume that physical distance between households does not affect the probability of casual contact. If a larger geographical area were to be modeled, this assumption would likely need to be modified since casual contacts are likely to occur in the same locality. The numbers of contacts in our model were chosen arbitrarily due to a lack of data and could be varied. Our simulations suggest that for a medium transmission rate (defined in Section 4.1), decreasing the number of casual contacts to 1–10 results in a roughly 20% decrease in incidence, while increasing the number of casual contacts to 50–100 results in a roughly 40% increase in incidence (data not shown).

The contact network generated from the synthetic population dataset for Washtenaw County produces a qualitatively similar age-based mixing pattern to that estimated for the US in a previous study by Prem et al. (2017). Most contacts occur between individuals of similar ages and the highest rates of contact are associated with teens and young adults. Since Washtenaw County contains a large university, there is a higher rate of contact among people in their early 20s than is predicted for the national average. For a graphical comparison of the age-based mixing patterns generated by our model and by Prem et al. (2017), please see our website at http://malthus.micro.med.umich.edu/synthetic/.

In the following virtual experiments, we simulate the spread of disease over a period of two years. For simplicity, and because we simulate over a relatively short length of time, we assume that the network is static; that is, the population and contact patterns do not change throughout the course of simulation. Thus, we do not include population dynamics such as births, deaths, or transitions between households, workplaces, schools, and group quarters. For simulations over several years or more, a realistic contact network would include these features and be dynamic to account for such changes over time.

#### 3.1.2 Model pathways and parameters

The NBM consists of many nodes (depending on the population size of the county), representing individual people in a population, each with its own demographic properties. The model structure and parameters are analogous to those of the continuous ODE and age-structured PDE model that we previously developed (Renardy and Kirschner, 2019; Guzzetta et al., 2011). Here, however, we do not track different types of active infection (e.g., reactivated vs primary infection) separately since these often cannot be distinguished in clinical practice and may not have a great impact during the short time frame we study here. At each time step in the simulation, agents are at one of these mutually exclusive disease states: susceptible (*S*), exposed (*E*), latent infection (*L*), active infection (*I*), and secondary exposures (*E*_*s*_). Agents can transition to another disease state via natural disease progression, treatment, or interaction with other agents.

If a susceptible agent comes into contact with an infected agent, the susceptible agent may become exposed. We assume that the probability of exposure is proportional to both the infected agent’s infectivity, which may correspond to severity of clinical symptoms, and the contact weight between the two agents. Contact weights between agents are determined by the population network structure discussed in Section 3.1.1. The exposed agent may 1) resolve the infection and move back to susceptible, 2) develop a latent infection, or 3) develop a primary infection. Agents with a latent infection may develop an active infection through 1) endogenous reactivation or 2) exogenous reinfection from secondary exposure. Agents with an active infection may “recover” from active infection through treatment and return to a latent state. We assume that these individuals return to a latent state rather than fully recovered or susceptible since it is not clear whether treatment kills all bacteria within a host, and treated individuals have been observed to spontaneously relapse (Gomez and McKinney, 2004; van Rie et al., 1999; Weis et al., 1994; Aktogu et al., 1996). At each time step, agents transition between states according to probabilities determined by the model parameters and contact weights. The model structure and pathways are summarized in Figure 1. Parameter values are taken primarily from Renardy and Kirschner (2019), where model parameters were calibrated to data for the US population. Model parameter values are summarized in Table 1. Rate parameters are given per year. To translate a per-year rate *r* to a probability *p* per time step, we use the formula 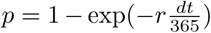, where the time step *dt* is measured in days.

**Table 1.** Values of model parameters taken from the literature. These parameter values are for the US population as a whole; we assume the same parameter values apply to Washtenaw County, MI.

| Parameter | Value | Units | References |
| --- | --- | --- | --- |
| Progression from exposure | 0.27672 | /yr | Renardy and Kirschner (2019); Guzzetta et al. (2011) |
| Probability of active disease upon infection (child) | 0.023375 |  | Renardy and Kirschner (2019); Guzzetta et al. (2011); Vynnycky and Fine (1997) |
| Probability of active disease upon infection (adult) | 0.23181 |  | Renardy and Kirschner (2019); Guzzetta et al. (2011); Vynnycky and Fine (1997) |
| Clearance of first infections | 0.73242 |  | Renardy and Kirschner (2019); Guzzetta et al. (2011) |
| Protection from exogenous reinfection (child) | 0.087282 |  | Renardy and Kirschner (2019); Guzzetta et al. (2011); Vynnycky and Fine (1997) |
| Protection from exogenous reinfection (adult) | 0.33939 |  | Renardy and Kirschner (2019); Guzzetta et al. (2011); Vynnycky and Fine (1997) |
| Reactivation rate | $2.1225 \times 10^{-4}$ | /yr | Renardy and Kirschner (2019); Guzzetta et al. (2011); Shea et al. (2014) |
| Successful treatment rate | 2.1032 | /yr | Renardy and Kirschner (2019); Guzzetta et al. (2011) |
| Proportion of population that is latent | 0.05 |  | Miramontes et al. (2015); Mancuso et al. (2016) |

**Fig. 1.**
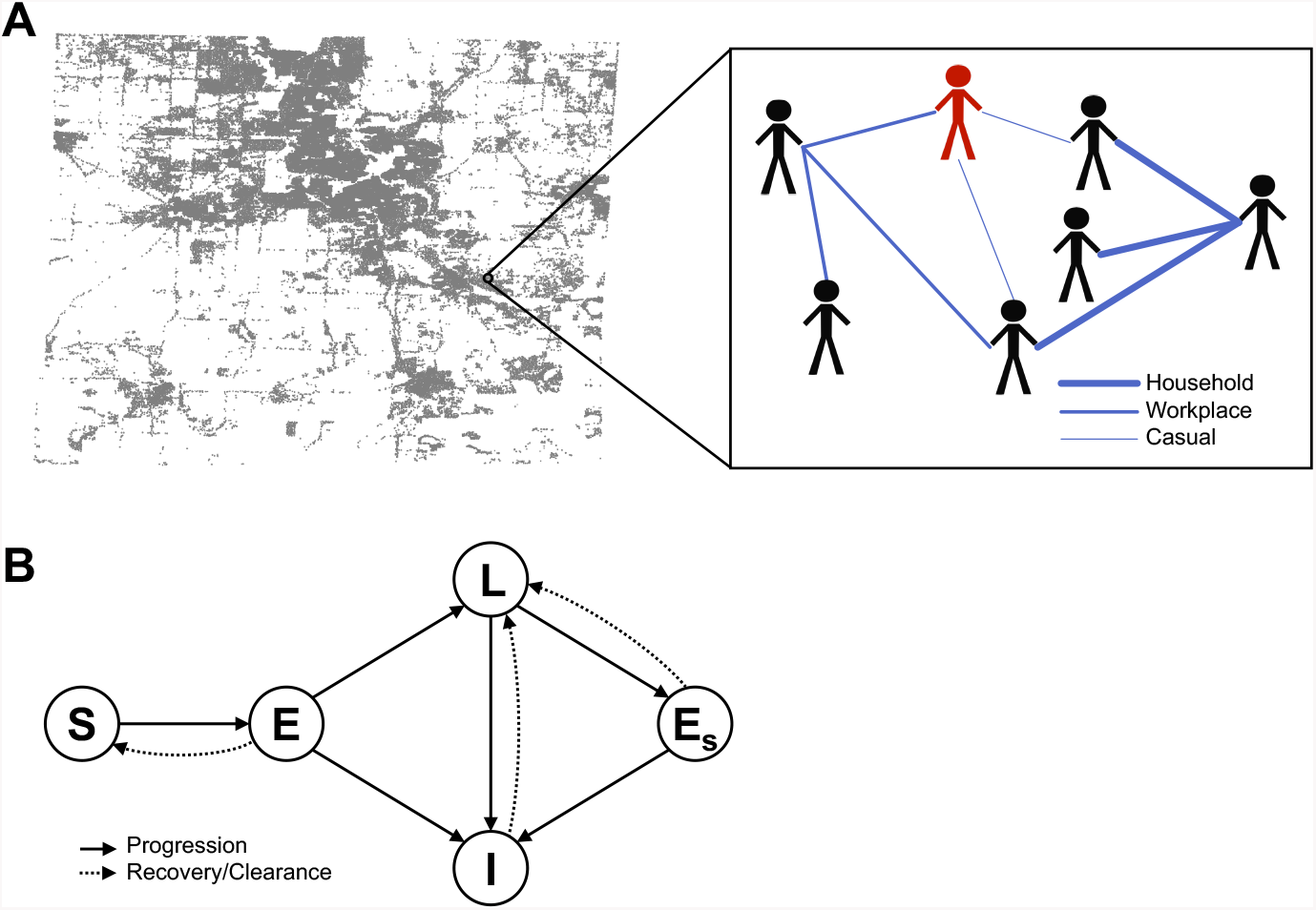
(A) Illustration of the population and network structure described in Section 3.1.1. Distribution of households in Washtenaw County and a hypothetical example of a small contact network with different types of contacts are shown. Thicker edges correspond to heavier contact weights. One individual is shown in red to indicate active infection. (B) Diagram showing all possible model pathways described in Section 3.1.2. *S* denotes susceptible, *E* denotes exposed, *L* denotes latent, *I* denotes infected, and *E*_*s*_ denotes secondary exposure.

Two model parameters are age dependent: probability of primary infection, and protection from reinfection. The functional forms for these parameters are identical to those used in Guzzetta et al. (2011) and Renardy and Kirschner (2019), and are given below as functions of age, where *p*(*a*) represents the probability of primary infection at age *a* and *σ*(*a*) represents the protection from reinfection at age *a*.

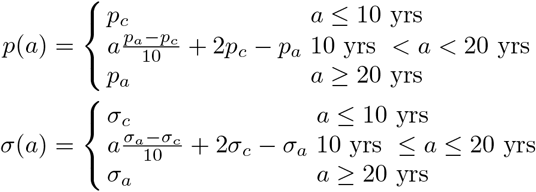

Here, *p*_*c*_ and *σ*_*c*_ represent the parameter values for children (under 10) and *p*_*a*_ and *σ*_*a*_ represent the parameter values for adults (over 20).

#### 3.1.3 Model initialization

The NBM is initialized by randomly infecting a single individual. Further, we assume that 5% of the population, chosen at random, carry a latent infection; this estimate is based on interferon gamma release assay (IGRA) blood test data from the National Health and Nutrition Examination Survey (NHANES), which studied a representative sample of the civilian, non-institutionalized US population (Miramontes et al., 2015; Mancuso et al., 2016). All other individuals are assumed to be susceptible.

#### 3.1.4 Implementation

The model is implemented in Matlab R2018b. Simulation runs were performed on a laptop computer with a 3.1 GHz Intel Core i7 processor and 16 GB 2133 MHz LPDDR3 RAM. In this computing environment, a single simulation over a 2-year period takes approximately 5 minutes of CPU time to complete. The Matlab code is provided on our website at http://malthus.micro.med.umich.edu/synthetic/.

### 3.2 Uncertainty and sensitivity analysis

To explore model behavior throughout the parameter space, we use Latin hypercube sampling (LHS) (McKay et al., 1979). This allows us to vary multiple parameters simultaneously within defined ranges using a uniform distribution. Since the network-based model has stochastic effects as well as randomized initialization, we simulate ten replicates for each parameter set as we have done previously (Marino et al., 2008).

We perform global sensitivity analysis using partial rank correlation coefficients (PRCC) to quantify the sensitivity of model outcomes to input parameters. The LHS/PRCC methodology for different types of model formulations is described in detail in Marino et al. (2008). This provides an estimate of the <u>epistemic</u> uncertainty in the model that results from unknown parameter values. PRCCs that are large in absolute value indicate sensitivity of the model output to input parameters, meaning that variation in parameter values result in significant variation in model output. PRCCs close to zero indicate non-sensitivity.

To quantify the <u>aleatory</u> uncertainty of the model, i.e., uncertainty that arises from stochasticity and random effects, we consider the <u>coefficient of variation</u> (CoV) of model outputs among multiple replicates for each parameter set. The coefficient of variation is a dimensionless quantity defined as the ratio of the standard deviation to the mean, which represents the relative variability of model outputs (Everitt, 1998).

## 4 Results

### 4.1 Effects of transmission rate and contact weights in a static network

One of the benefits of using a network-based model is that transmission events can be traced and the type(s) of contact that lead to a transmission event can be determined. To explore the effects of different types of contacts on model dynamics, we performed LHS and sensitivity analyses for the different types of contact weights. Contact weights were normalized so that the contact weight for household contacts is equal to 1. Since there are no data available for the values of different contact weights in Washtenaw County, we allowed these contact weights to vary within broad ranges. Other contact weights were allowed to vary from 0.5–1 for workplace and school contacts and 0.01–0.2 for casual contacts. We assume that group quarters contacts are equivalent to household contacts. All types of contact, including casual contacts, are static, meaning that they persist over the full time-course of the simulation.

Repeat and prolonged exposures between infected and uninfected individuals are thought to be key routes for transmission for TB (Sepkowitz, 1996; Dobler et al., 2016); thus, household transmissions are thought to contribute significantly to epidemiological dynamics. However, the household transmission rate for TB in the US is unknown. Thus, we consider three different cases to explore the effects of transmission rates that are low (0.75/yr), medium (7.5/yr), and high transmission (75/yr). We note that these represent low, medium, and high transmission rates <u>in the US</u> and that transmission rates in higher incidence countries are likely to be significantly higher. We fixed the remaining model parameters at the values that we fit to the US population for an age-structure PDE model in Renardy and Kirschner (2019). For each choice of transmission rate, we performed LHS to obtain 100 uniformly distributed contact weight parameter sets in the ranges described above, given in Table 2. For each parameter set that is sampled, ten replicate simulations are performed to account for random effects during initialization and simulation. We believe this sample size is sufficient since results did not substantially change upon repetition of with new random samples (data not shown). In these simulations, we use a fixed time step of one week and simulate over the course of two years.

**Table 2.** Parameter values and ranges for parameters that are newly created for this model or whose values are different from those used in previous models.

| Parameter | Value/range | Unit |
| --- | --- | --- |
| Transmission rate | 0.75 (low), 7.5 (medium), 75 (high) | per year |
| Household contact weight | 1 | dimensionless |
| Workplace contact weight | 0.5–1 | dimensionless |
| School contact weight | 0.5–1 | dimensionless |
| Group quarter contact weight | 1 | dimensionless |
| Casual contact weight | 0.01–0.2 | dimensionless |

On average, over a simulated time of two years, NBM simulations predict that the average incidence rates are 1.2, 2.3, and 6.5 per 100,000 per year for the low, medium, and high transmission cases, respectively. For reference, the actual average incidence of TB in Washtenaw County, MI over the past ten years has been 2.1 per 100,000 per year (Washtenaw County Health Department, 2019), aligning best with the prediction of the medium transmission case. In our simulations, we find that in the low transmission rate case, the vast majority of active infections after two years are due to reactivation of latent infections. Reactivation plays a much smaller role in the cases of medium and high transmission rates. In the medium transmission rate case, workplace transmission is responsible for the most infections, followed by reactivation and then by casual contacts. In the high transmission rate case, casual contact transmission is responsible for the most infections, followed by workplace transmission, with reactivation playing only a minor role. The distributions of active infections by source of infection are shown in Figure 2 (top row).

**Fig. 2.**
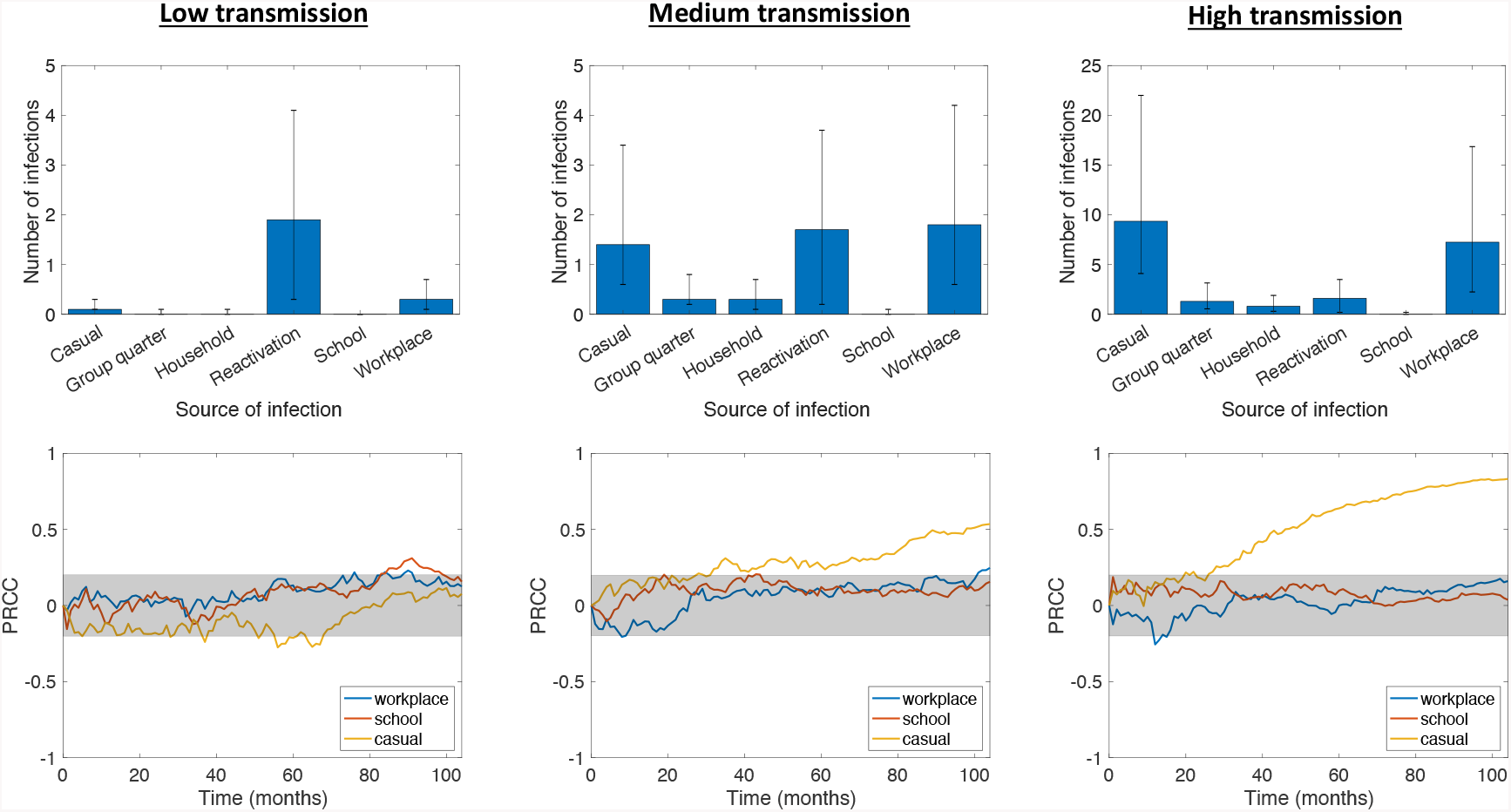
Top row: Distribution of average number of active infections at *t* = 2 years by source of infection. Bar heights represent median values, while error bars represent the 25th and 75th percentiles. Bottom row: Sensitivities of active TB prevalence over time to different types of contact weights, measured via partial rank correlation coefficients (see Section 3.2, −1 *≤ PRCC ≤*1). The gray shaded area indicates sensitivity values that are not statistically significant using a p-value of 0.05. Contact weights were randomly sampled via LHS with low, medium, and high transmission rates; other model parameters were fixed at the values in Renardy and Kirschner (2019).

The higher number of casual infections in the high transmission case likely occurs because, in our model, individuals tend to have more casual contacts than workplace contacts. One reason for this is that the upper bound on the number of workplace contacts is equal to the upper bound on the number of casual contacts, and thus, generally, the number of workplace contacts does not significantly exceed the number of casual contacts. Further, only about 50% of the population belongs to a workplace and many workplaces are quite small. In the high transmission case, 30% of runs had a median of fewer than 10 workplace contacts among infected individuals. By contrast, every individual is assigned at least 10 (and at most 50) casual contacts. Thus, while the upper bounds for workplace and casual contacts are the same, a significant number of individuals have fewer workplace contacts than casual contacts. This effect is not observed at lower transmission rates because the number of infected individuals and the transmission rate among casual contacts are sufficiently low.

We note that in all three cases, the proportion of active infections resulting from household and group quarters transmissions is very low despite being associated with the largest contact weights. While this may seem counterintuitive, similar phenomena have been observed in several epidemiological studies of TB in a variety of settings. In England, for example, a retrospective study of TB cases from 2010 to 2012 found that only 3.9% were due to recent household transmission (Lalor et al., 2017). In suburban South Africa, a much higher incidence setting than is considered here, analysis of TB cases between 1993 and 1998 revealed that only 19% of transmission in the community took place within households (Verver et al., 2004). A low proportion of household transmission in South Africa was also supported by a previous study in a rural setting (Wilkinson et al., 1997). In the context of Washtenaw County, the low proportion of household-related active infections is likely due to the small size of households. The median household size in our synthetic population is 2, and 31% of households have only one member.

There were also very few cases of active infections caused by school transmission. This is primarily the result of children being less likely to develop active infection. Studies in England and Wales have shown that children have a significantly lower probability of active disease upon infection (Vynnycky and Fine, 1997), which is reflected in our model parameters (see Table 1). This reduces the number of active infections resulting from school transmission in two ways. First, due to age demographics, there are relatively few active infections that occur in individuals that belong to schools. In the high transmission case, across all simulations, only 9.1% of actively infected individuals belonged to a school, whereas 65.8% of actively infected individuals belonged to a workplace. Second, transmissions that occur within schools are more likely to lead to latent disease than transmissions that occur within workplaces.

We performed sensitivity analysis using partial rank correlation coefficients (PRCC) to evaluate the effects of workplace, school, and casual contact weights on TB prevalence over time. The sensitivities over time are shown in Figure 2 (bottom row). In the low transmission case, we find that TB prevalence is not significantly sensitive to any of the contact weights. In the medium and high transmission cases, we find that prevalence is sensitive to the casual contact weight but not to the school or workplace contact weights. This implies that as transmission rate increases within a population, sensitivity of TB prevalence to the casual contact weight also increases. Intuition suggests that this follows from a combination of 1) casual contacts allowing for wider disease spread by enabling transmission between different workplaces and households, and 2) higher transmission rates making transmission by casual contact more likely. Increased connectivity between households and workplaces leads not only a more connected contact network, but also potentially greater heterogeneity among the population that could be exposed to disease. Both of these factors have been shown to contribute to increased likelihood of disease invasion and persistence (Dushoff and Levin, 1995; Keeling, 2005; Gupta et al., 1989).

The relative frequency of reactivated infections in the low-transmission network model is consistent with a recent study that analyzed 26,586 genotyped TB cases in the US from 2011–2014 and revealed that, among the 49 US states included, the proportion of infections attributable to recent transmission varied from 0% to 51% (Yuen et al., 2016). In the low-transmission network model of Washtenaw County, 21% of TB cases after two years were due to recent transmission, on average. In the medium transmission case, 72% were due to recent transmission. During the 2011–2014 time period, average incidence in Washtenaw County, MI was 1.8 per 100,000, which is halfway between the simulated low and medium transmission cases (Washtenaw County Health Department, 2019).

### 4.2 Random effects lead to challenges in model calibration

Computational models are often calibrated to match available experimental data, which can be complicated by aleatory uncertainty in model outputs. In our NBM, model initialization is random and transmission and reactivation events occur stochastically; thus, there is significant variation in model outputs among replicates even using the same set of values for model parameters. To quantify this variation in each of the low, medium, and high transmission cases, we compute a CoV of the number of infected individuals across replicates for each set of parameter values in the LHS (100 samples) and at each time point in the simulation (2 years *×* 52 weeks/year = 104 time points). The distributions of these coefficients for the three cases is shown in Figure 3A. We find that the low, medium, and high transmission cases are nearly identical. The average CoV is 0.75 for the low and medium transmission cases, and 0.8 for the high transmission case. Thus, regardless of transmission rate, the standard deviation across replicates on average for a single parameter set is roughly 3/4 of the mean.

**Fig. 3.**
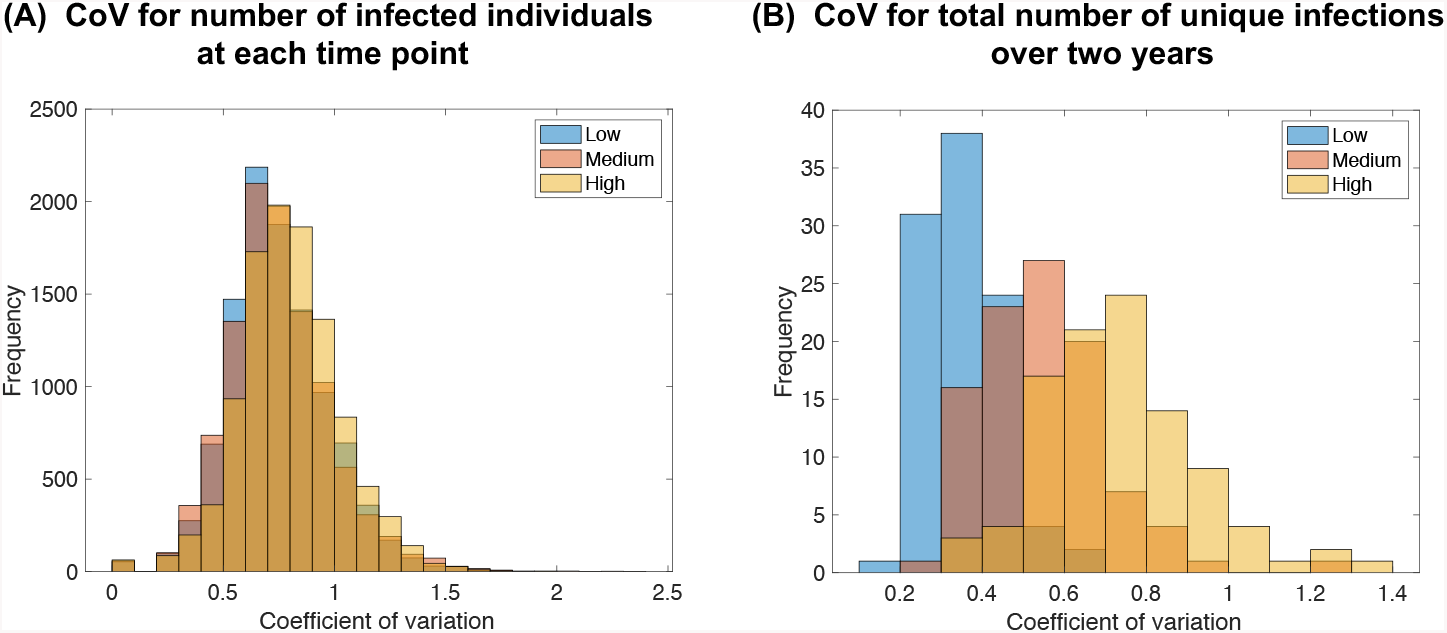
Variation in model outputs quantified by coefficients of variation. We simulated 10 replicates for each parameter set in the LHS (100 samples). (A) Coefficients for the number of infected individuals at each time point (104 time points). (B) Coefficients for the total number of unique infections over the entire two-year period.

Variation in model outputs could be reduced by more informed initial conditions, such as by using socio-demographic information to place initial active and latent infections in a way that is consistent with the true population. However, this requires access to data that is often nonexistent (particular for latent infections) or not publicly available. Further, our computational experiments suggest that using a fixed initial condition results in only a minimal reduction in variation. For the medium transmission case, using a fixed initial condition for all 10 replicates of each parameter set led to an average CoV of 0.73, which is not a meaningful decrease from the CoV using random initializations. Thus, the majority of variation in model output results from stochasticity of the model and not from randomized initial conditions.

Much of the variation discussed above is due to temporal variation in the number of infections, since the CoV was evaluated at every time step. If we instead consider the total number of unique infections over the entire two-year period, we find that the CoVs are reduced to 0.35, 0.56, and 0.77 in the low, medium, and high transmission cases, respectively; the distributions are shown in Figure 3B. Thus, we observe that there is less relative variation in the total number of unique infections over a period of two years at lower transmission rates, and this variation is generally lower than the variation in the number of infected individuals at any given time. This implies that output variation can be reduced by considering summary statistics over long time periods, as would be expected.

Still, stochastic effects could lead to significantly different model predictions if model parameters are calibrated using available prevalence or incidence data based on average model behavior. This issue is further compounded by possible unidentifiability of model parameters even in the absence of stochastic effects. This means that one may not be able to uniquely estimate parameter values from observable data such as prevalence and incidence. Issues of parameter unidentifiability have been explored in the context of continuous epidemic models and have been shown to lead to incorrect predictions for the effects of interventions (Kao and Eisenberg, 2018). This presents serious challenges in model calibration and the interpretation of model-based predictions.

### 4.3 Localization of simulated infections

One major benefit of using a spatial NBM over non-spatial models such as ODE or age-structured models is the ability to obtain spatial information about epidemiological dynamics. Spatial information at a sub-county level for TB in the US is often unavailable due to patient privacy concerns. The Report of Verified Case of Tuberculosis (RVCT) Form, which is used for national TB surveillance, includes information on city, county, and zip code (CDC, Division of Tuberculosis Elimination, 2009); however, access to this information requires approval from the state or local health department. Thus, in cases where this information cannot be obtained, modeling can yield insights into probable spatial patterns of infections. In cases where spatial information is available, this could be used to validate a model.

To quantify the spatial distribution of simulated infections, we compute the average pairwise distances between infected individuals at the end of the two-year simulation for each parameter set in the LHS (sample size = 10 replications *×*100 parameter sets = 1000). An individual’s location is defined by the location of their household or group quarter. Among the simulations for which more than one individual was infected at the end of two years, the pairwise distances averaged 10.7 *±* 6.0 miles for the low transmission case, 10.4 *±*4.6 miles for the medium transmission case, and 10.3 *±* 3.1 miles for the high transmission case. For comparison, we considered 10,000 samples where a random number (up to 20) of individuals are uniformly and randomly chosen from the population. Among the randomly selected individuals, the pairwise distance averages 10.8 *±*3.1 miles.

The difference between the low and medium transmission cases and the random sample is not statistically significant. This is expected for the low transmission case since most active infections in this case are due to reactivation of latent infection and the latent population is randomly distributed. However, for the high transmission case, the pairwise distances between infected individuals are smaller than if infections were purely random; while these differences are not particularly large, they are statistically significant (*p <* 0.001, significance determined using a *t*-test). This implies that although workplaces and casual contacts play a much larger role than households in recent transmissions, infections are still localized based on household locations. Spatial distributions for representative samples from the three transmission cases as well as randomly sampled individuals are shown in Figure 4.

**Fig. 4.**
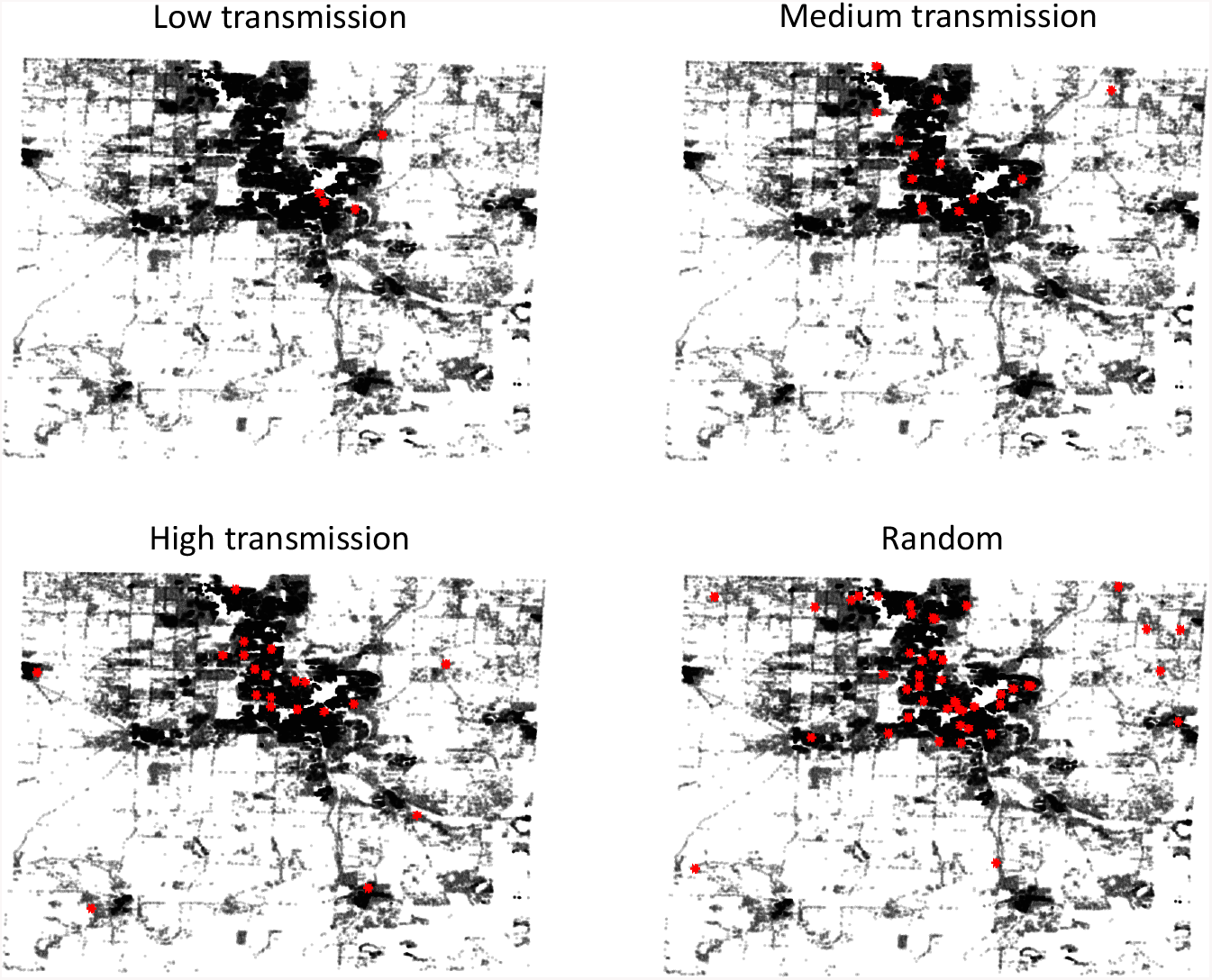
Simulated maps showing locations of all households in Washtenaw County (black) and the household locations of infected individuals (red) at the end of a two-year simulation for representative samples from the low, medium, and high transmission cases and for uniformly random individuals as a comparison.

The observed localization by household location in our simulations is likely due to individuals who live close together being more likely to work close together. Simulations also showed statistically significant localization by workplace locations among those infected individuals who belonged to a workplace. However, we focus here on the spatial distribution of infections by households as all individuals in the synthetic population belong to a household or group quarter, and all households and group quarters are located within Washtenaw County. Further, household location is more likely to be reported in epidemiological data than workplace location.

## 5 Discussion

The purpose of this paper is to establish and demonstrate a network-based model for TB epidemiology, which utilizes publicly available synthetic datasets to create a realistic contact network and assign socio-demographic attributes to the population. We do not calibrate the model to epidemiological data within Washtenaw County, and thus we make no specific predictions about the spread of TB. Rather, we use randomization and parameter exploration to study general model behavior. Washtenaw County was chosen as a test population due to its proximity to University of Michigan, where this research was conducted.

There are many benefits to using a discrete network-based model over other model formulations such as ODE and PDE models. Most notably, discrete models such as the one considered here allow for host heterogeneity to be explicitly accounted for. Using a network structure within a discrete model allows for establishing contact patterns within a population without needing to explicitly model movement on a spatial domain, which can become expensive when the number of individuals is large. Synthetic population datasets are an excellent tool to provide realistic contact networks, socio-demographic heterogeneity in the relevant population, and explicit geospatial data.

Using these synthetic population datasets allows us to consider different types of contacts and their influence on TB epidemiology as well as the spatial distribution of infections. For example, we found that the majority of active transmission in our model occurs in workplaces and through casual contacts, while household, group quarter, and school contacts play a minor role. This is primarily due to individuals in our synthetic population that tend to have more workplace and casual contacts than any other type of contact, and school-related contacts do not significantly contribute to transmission due to a decreased occurrence of primary infection in children. Further, we have found that at transmission rates large enough to create a substantial number of active transmission events, disease prevalence is sensitive to the contact weight for casual contacts, where the contact weight represents the duration of contact, and is not sensitive to any other contact weights.

As with any model, many of the results presented here inherently depend on the choices of ranges for contact weights as well as other parameters, and results may differ for different parameter values. Here, our focus is on the model framework and exploration of model outcomes rather than specific predictions about TB epidemics. Thus, we have allowed contact weights to vary in broad ranges, but we have not calibrated the model to match specific epidemiological data as that was not the goal of this work. To produce reliable model-based predictions of epidemic dynamics or intervention efficacy, parameter estimation will be crucial; however, this is beyond the scope of this paper, but will be addressed in future work. Since disease incidence is not sensitive to most of the contact weight parameters, these parameters likely cannot be estimated from incidence data. Thus, to calibrate these parameters, more detailed epidemiological data would be needed. For example, demographic data for infected individuals could be used to create more informed initial conditions as well as to provide additional data for estimating parameters. Contact data such as that from Mossong et al. (2008) and Prem et al. (2017) could be used to calibrate contact weights; however, these data are at the national scale and may not accurately reflect county level rates.

One potential disadvantage to the use of a stochastic discrete model is the uncertainty introduced by random effects. Computational experiments suggest that random effects in our model lead to a significant amount of variation in the predicted number of infections even for constant parameter values, which could lead to challenges in model calibration and interpreting model-based predictions. However, this variation may also be seen as an advantage since the real-world system is likely affected by substantial randomness and this model allows us to explore many different possible outcomes.

Our model can be used to explore epidemiological dynamics at the county level to design intervention strategies for specific local populations. For example, effectiveness and cost of interventions may differ dramatically between urban and rural counties even if they are geographically close; thus, considering these localities separately may lead to more effective interventions. Comparison of epidemiological dynamics across localities may also yield insights into differences in driving factors for disease spread, transmission dynamics, and risk groups. Further, some counties within the US may be demographically similar to developing countries; thus, county-level modeling can provide insight for how interventions could work at a larger scale for high-incidence settings. Intervention strategies can be expensive and cumbersome to implement in practice, and they may have unintended consequences; thus, mathematical and computational modeling are critical tools for aiding in the discovery, application, and evaluation of intervention strategies. Previous modeling efforts have addressed TB treatment and vaccination effectiveness (Abu-Raddad et al., 2009; Castillo-Chavez and Feng, 1998; Knight et al., 2014; Lietman and Blower, 2000; Renardy and Kirschner, 2019; Ziv et al., 2004) and other interventions such as contact tracing (Kasaie et al., 2014; Tian et al., 2013).

Since TB can progress and persist over long periods of time, epidemiological studies may require modeling disease spread over several years or even decades. In such studies, the population and contact network should be dynamic to account for births, deaths, and transitions between households, workplaces, schools, and group quarters. This was not done here since we considered a time period of only two years for simplicity. In future studies, this model can also be improved by obtaining better-informed initial conditions and parameter estimates using socio-demographic data of the real infected population. At the county level, since TB is a low incidence disease in the US, accessing this type of data will require special permissions due to issues of patient privacy. Better interactions of modelers with local and national health agencies will likely aid in advancing our understanding and ultimately interventions that can eliminate TB.

## Data Availability

Relevant code and datasets are provided on our website at http://malthus.micro.med.umich.edu/synthetic/.

## Acknowledgements

This research was supported by NIH grants R01AI123093 and U01 HL131072 awarded to DEK. The 2010 U.S. Synthetic Population database was created by RTI International, which is funded by the National Institutes of General Medical Sciences (NIGMS).

